# Detecting COVID-19 infection hotspots in England using large-scale self-reported data from a mobile application: a prospective, observational study

**DOI:** 10.1101/2020.10.26.20219659

**Authors:** Thomas Varsavsky, Mark S. Graham, Liane S. Canas, Sajaysurya Ganesh, Joan Capdevila Pujol, Carole H. Sudre, Benjamin Murray, Marc Modat, M. Jorge Cardoso, Christina M. Astley, David A Drew, Long H. Nguyen, Tove Fall, Maria F Gomez, Paul W. Franks, Andrew T. Chan, Richard Davies, Jonathan Wolf, Claire J. Steves, Tim D. Spector, Sebastien Ourselin

**Affiliations:** School of Biomedical Engineering & Imaging Sciences, King’s College London, London, UK; Zoe Global Limited, London, UK; MRC Unit for Lifelong Health and Ageing, Department of Population Science and Experimental Medicine, University College London, UK; Centre for Medical Image Computing, Department of Computer Science, University College London, UK; Division of Endocrinology and Computational Epidemiology, Boston Children’s Hospital, Harvard Medical School, USA; Clinical and Translational Epidemiology Unit, Massachusetts General Hospital, MA, USA; Department of Medical Sciences, Uppsala University, Uppsala, Sweden; Department of Clinical Sciences, Lund University Diabetes Centre, Sweden; Genetic and Molecular Epidemiology Unit, Lund University Diabetes Centre, Sweden; .Department of Twin Research and Genetic Epidemiology, King’s College London, London, UK

## Abstract

**Background:** As many countries seek to slow the spread of COVID-19 without reimposing national restrictions, it has become important to track the disease at a local level to identify areas in need of targeted intervention.

**Methods:** We performed modelling on longitudinal, self-reported data from users of the COVID Symptom Study app in England between 24 March and 29 September, 2020. Combining a symptom-based predictive model for COVID-19 positivity and RT-PCR tests provided by the Department of Health we were able to estimate disease incidence, prevalence and effective reproduction number. Geographically granular estimates were used to highlight regions with rapidly increasing case numbers, or hotspots.

**Findings:** More than 2.8 million app users in England provided 120 million daily reports of their symptoms, and recorded the results of 170,000 PCR tests. On a national level our estimates of incidence and prevalence showed similar sensitivity to changes as two national community surveys: the ONS and REACT-1 studies. On 28 September 2020 we estimated 15,841 (95% CI 14,023-17,885) daily cases, a prevalence of 0.53% (95% CI 0.45-0.60), and R(t) of 1.17 (95% credible interval 1.15-1.19) in England. On a geographically granular level, on 28 September 2020 we detected 15 of the 20 regions with highest incidence according to Government test data, with indications that our method may be able to detect rapid case increases in regions where Government testing provision is more limited.

**Interpretation:** Self-reported data from mobile applications can provide an agile resource to inform policymakers during a fast-moving pandemic, serving as an independent and complementary resource to more traditional instruments for disease surveillance.

**Funding:** Zoe Global Limited, Department of Health, Wellcome Trust, EPSRC, NIHR, MRC, Alzheimer’s Society.

**Research in context:** *Evidence before this study:* To identify instances of the use of digital tools to perform COVID-19 surveillance, we searched PubMed for peer-reviewed articles between 1 January and 14 October 2020, using the keywords COVID-19 AND ((mobile application) OR (web tool) OR (digital survey)). Of the 382 results, we found eight that utilised user-reported data to ascertain a user’s COVID-19 status. Of these, none sought to provide disease surveillance on a national level, or to compare these predictions to other tools to ascertain their accuracy. Furthermore, none of these papers sought to use their data to highlight geographical areas of concern.

*Added value of this study:* To our knowledge, we provide the first demonstration of mobile technology to provide national-level disease surveillance. Using over 120 million reports from more than 2.8 million users across England, we estimate incidence, prevalence, and the effective reproduction number. We compare these estimates to those from national community surveys to understand the effectiveness of these digital tools. Furthermore, we demonstrate the large number of users can be used to provide disease surveillance with high geographical granularity, potentially providing a valuable source of information for policymakers seeking to understand the spread of the disease.

*Implications of all the available evidence:* Our findings suggest that mobile technology can be used to provide real-time data on the national and local state of the pandemic, enabling policymakers to make informed decisions in a fast-moving pandemic.

## Introduction

The COVID-19 pandemic caused many countries to impose strict restrictions on their citizen’s mobility and behaviour to curb the rapid spread of disease, often termed ‘lockdowns’. Since relaxing these restrictions, many countries sought to avoid their re-imposition through combinations of non-pharmaceutical interventions^1^ and test-and-trace systems. Despite these efforts, many countries have experienced rises in infection since opening and have often re-imposed either regional^2^ or national lockdowns. Regional lockdowns aim to contain the disease whilst minimising the severe economic impact of national lockdowns.

The effectiveness of regional interventions depend on the early detection of so-called infection hotspots^3^. Large-scale, population-based testing can indicate regional hotspots, but at the cost of a delay between testing and actionable results. Moreover, accurately identifying changes in the infection rate requires sufficient testing coverage of a given population^4^, which can be costly and requires significant testing capacity. Regional variation in testing access may hamper the ability of public health bodies to detect rapid changes in infection rate. There is a high unmet need for tools and methods that can facilitate the timely and cost-effective identification of infection hotspots to enable policymakers to act with minimal delay^5^.

In this work we aim to use self-reported population-wide data, obtained from a mobile application (the COVID Symptom Study app), combined with targeted PCR testing to provide geographical estimates of disease prevalence and incidence. We further show how these estimates can be used to provide timely identification of infection hotspots.

## Methods

### Application data

Data were collected using the COVID Symptom Study app, developed by Zoe Global Ltd with input from King’s College London, the Massachusetts General Hospital, and Lund and Uppsala Universities. The app guides participants through a set of enrolment questions, establishing baseline demographic information. Users are asked to record each day whether they feel physically normal, and if not, to log any symptoms experienced, and to keep a record of any COVID-19 tests and their results (appendix pp 3-6). More details about the app can be found in^6^, which also contains a preliminary demonstration of how symptom data may be used to estimate prevalence. In England, the Department of Health and Social Care (DHSC) allocated COVID-19 tests to users of the Study app, beginning on 28 April. Users who logged as healthy at least once in a nine day period and then reported any symptom (which we term ‘newly sick’) were sent invitations to book a test through the DHSC’s national testing programme, and asked to record the result of the test in the app (Fig S6). This work only includes app users living in England, as some of the methods described make use of this England-specific testing capacity. We include responses logged between 24 March and 29 September. Our study was approved by King’s College London ethics committee (REMAS ID 18210, LRS-19/20-18210).

### External datasets

Three datasets were used for validation of our models: the Office for National Statistics (ONS) Community Infection Survey, the Real-time Assessment of Community Transmission (REACT-1) study, and Government testing data. The ONS survey^7^ is a longitudinal survey of individuals selected to be a representative sample of private households (excluding e.g. care homes, student accommodation). Individuals are supervised while they self-administer nose and throat swabs. The results give estimates of prevalence and incidence over time, with the first estimates being on 30 April. In September they recorded 151,000 participants who had provided at least one swab result. Data is released weekly, with each release covering the period 7-14 days before the release date. The REACT-1 study is a cross-sectional community survey, relying on self-administered swab tests from a representative sample of the population in England^8,9^, ranging between 120,000 and 175,000 people in each round of data collection. Data releases are intermittent and cover periods of several weeks. The Government swab-test data is made up of two ‘Pillars’ of testing: Pillar 1 covers those with clinical need and healthcare workers, and Pillar 2 testing covers the wider population who meet Government guidelines for testing^10^. The ONS and REACT-1 surveys were used to compare our national estimates of incidence and prevalence. The Government testing data was used to validate our geographically granular list of hotspots.

### Incidence

Incidence is calculated using the invited swab tests reported in the app. We took 14-day averages starting on 12 May to calculate the percentage of positive tests amongst newly sick users per National Health Service (NHS) region in England, and combined this with the proportion of users who report as newly sick in that region to produce the probability that a randomly selected person in that region is infected with COVID-19 on a given day (appendix p 2). We multiply this probability by the population of each region to produce our swab-based incidence estimates, which we term I_S_. Incidence values are released daily, with a four-day reporting lag.

### Prevalence

We describe two methods for estimating prevalence. The first (symptom-based) primarily makes use of self-reported symptoms and a predictive, symptom-based model for COVID-19. The second (symptom and swab based) seeks to further integrate the information from swab test results collected in the app.

#### Symptom-based

The symptom-based approach uses a previously validated logistic regression model^11^ to predict whether a user is COVID-19 positive or not, based on their reported symptoms (appendix p 2). For a given day, each user’s most recent symptom report from the previous seven days is used for prediction. If a user reports a positive COVID-19 test in that 7-day period, its result is used to override the user’s symptom-based estimate. The proportion of positive users are used to estimate prevalence. A user that is predicted COVID-19 positive for more than 30 days is considered long-term sick and no longer infectious, and removed from the calculation. We sought to extrapolate these prevalence estimates to the general population. As noted in previous work^12^ there is a disparity in COVID-19 prevalence between regions of higher Index of Multiple Deprivation (IMD), a measure of the relative deprivation of geographical regions^13^, and those of lower IMD. We stratify users by Upper Tier Local Authority (UTLA), IMD tertile, and age bands (in decades), and predict percentage prevalence per strata. We then multiply our predicted percentage of positive cases per strata with the strata’s population size according to census data to estimate cases per strata. These are then summed to produce our population prevalence estimate. We term this prevalence P_A_. We examined sensitivity of P_A_ to health-seeking bias by calculating the quantity removing all users reporting sick at sign-up.

#### Symptom and swab based

To estimate prevalence using the swab data, we make use of our incidence estimates and the relationship

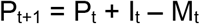

where M_t_ represents the number of patients that recover at time t and I_t_ the number of new COVID-19 cases at time t. We estimate M_t_ from our data (appendix p3).

These prevalence estimates make use of the swab results but lack geographical granularity, being per NHS region. We can increase the granularity by taking the symptom-based estimates, which are calculated per UTLA, and rescaling all the estimates that make up an NHS region such that the total prevalence across those UTLAs matches the per-NHS region prevalence we estimate. We term this hybrid method, making use of both symptom-reports and swab-tests, as P_H_. It is possible to produce granular incidence estimates by applying the model of recovery to these granular prevalence estimates: we term these estimates I_H_.

### Calculating R(t)

It is possible to retrieve R(t) from incidence rates by combining them with known values of the serial interval^14^. Briefly, we use the relationship

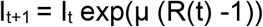

Where 1/µ is the serial interval. We model the system as a Poisson process and use Markov Chain Monte-Carlo (MCMC) to estimate R(t). In our probabilistic modelling we assumed that the serial interval was drawn from a Gamma distribution with α=6.0 and β=1.5 as in^15^. By sampling successive chains from the system we obtain a distribution over R(t), which allows us to report a median and 95% credible intervals. These estimates of uncertainty do not take into account the uncertainty in the estimate of incidence, which we found to be mostly systematic and smaller than the other forms of uncertainty modelled.

### Hotspot detection

A hotspot is defined as a sudden increase in the number of cases in a specific geographic region. We produce two rankings of UTLAs in England. The first ranks each by their estimated prevalence P_H._ This has the advantage of being pre-registered; a list of the top ten UTLAs according to P_H_ has been published online since 23 July. However, this does not allow the direct identification of areas of concern, i.e. areas with a large number of new cases, and so we report a second ranking using I_H_.

We compared our rankings to those obtained by ranking according to Government testing data. England contains 149 UTLAs, containing a mean of 370,000 people. We used the Government data to produce daily reference rankings of each UTLA, based on seven-day moving averages of daily cases per UTLA. We included all tests performed on a given day to produce the ranking for that day, even if that test took several days to have its result returned, to produce the most accurate ‘gold standard’ ranking we could. We used 7-day moving averages of P_H_ and I_H_ to produce our predicted rankings of each UTLA. We then evaluated these predictions against the historic reference using two metrics. The first, recall at 20, is the number of UTLAs in our top twenty that appear in the reference top twenty. The second, the normalised mean reciprocal rank at 20, measures the agreement between ranks of our top twenty list. We estimated uncertainty by drawing 100 samples of P_H_, I_H_, and the Government testing data for each UTLA and day, making use of errors calculated using the Wilson interval approximation for the Binomial distribution. These samples were ranked and metrics re-computed to produce 95% CIs for each metric.

### Statistical analysis

Power analysis found that 320,000 weekly active users are needed to detect an increase from 5,000 to 7,500 daily cases at p <0.05. All analysis was done with Python 3.7. ExeTera 0.2.7 was used for dataset processing and analysis.^16^ The COVID Symptom Study app is registered with ClinicalTrials.gov, NCT04331509.

### Role of the funding source

Zoe Global developed the app for data collection. The funders had no role in the study design, data collection, data analysis, data interpretation, or writing of the report. All authors had full access to all the data and the corresponding author had final responsibility for the decision to submit for publication.

## Results

From 24 March to 29 September, 4,644,227 participants signed up to use the app, of which 2,873,726 reported living in England. After excluding participants with invalid age information (6,230) and those without any daily assessments logged (26,422) we report 2,841,074 users that participated in this study. These participants completed 120,154,058 daily assessments, or 42.3 per user, with a median 16 logs (IQR 3 - 68), corresponding to logged data for 22.5% of the total possible person-logs over the study period. Compared to the population of England, we have fewer participants under 18 (6.9% compared with 22.5%) and fewer older than 65 (12.3% vs 18.4%), our participants are more female (61.2% vs 50.6%) and live in less deprived regions (46.5% live in the least deprived tercile of regions, compared with 33.3%). Between 28 April and 29 September, 851,250 invitations for swab tests were sent out. Of these 169,682 people reported a swab test on the app, of which 1,912 tested positive. In addition, users reported the results of 689,426 non-invited tests in the app, of which 25,663 were positive. 621,031 symptom reports were classified as COVID-19 positive by the symptom model.

**Table 1.** Characteristics of all app users in England that signed up between 24 March and 29 September, 2020.

|  |  | N | Percentage |
| --- | --- | --- | --- |
| **Users** |  | 2,841,074 | --- |
| **Daily reports** |  | 120,154,058 |  |
| **Age in years mean (std)** |  | 43.26 (17.27) | --- |
|  | ≤18 | 195,953/2,841,074 | 6.90% |
|  | 19 - 64 | 2,295,265/2,841,074 | 80.79% |
|  | ≥ 65 | 349,856/2,841,074 | 12.31% |
| **Female** |  | 1,739,929/2,841,074 | 61.24% |
| **Pre-conditions\*** | Kidney disease | 18,658/2,840,372 | 0.66% |
|  | Lung disease | 302,494/2,505,317 | 12.07% |
|  | Heart disease | 66,399/2,840,372 | 2.34% |
|  | Diabetes | 79,090/2,840,372 | 2.78% |
|  | Cancer** | 19,530/1,507,045 | 1.30% |
| **IMD** | Bottom tercile (most deprived) | 529,715/2,841,074 | 18.64% |
|  | Middle tercile | 989,543/2,841,074 | 34.83% |
|  | Upper tercile (least deprived) | 1,321,816/2,841,074 | 46.53% |
| **Smokers** |  | 275,822/2,841,074 | 9.71% |

\*Not all participants answered questions on preconditions. Numbers reported are absolute number of users that reported having the precondition, percentages are calculated amongst all participants that answered each precondition question.
\*\*Question asked from 29 March

Figure 1A compares England-wide incidence estimates I_S_ to Government testing data, and the ONS survey^17^. We include two estimates from the ONS: the official reports, released every week, and the results from time-series modelling. The reports represent the ONS best estimate at the time of release, whilst the times-series model can evolve and lead to revision of previous estimates in response to new data (see Figure S1 for an example of this). The Government figures are consistently lower than other estimates because they are not a representative figure for the population. To account for this, we looked at the number of people who reported classic symptoms (fever, loss of smell and persistent cough) for the first time between 7 July and 5 August and who did not get tested; we found the number to be 59%. We used this to scale the Government data by a factor of 2.5, our best estimate of the systematic undercounting of new cases.

**Figure 1.**
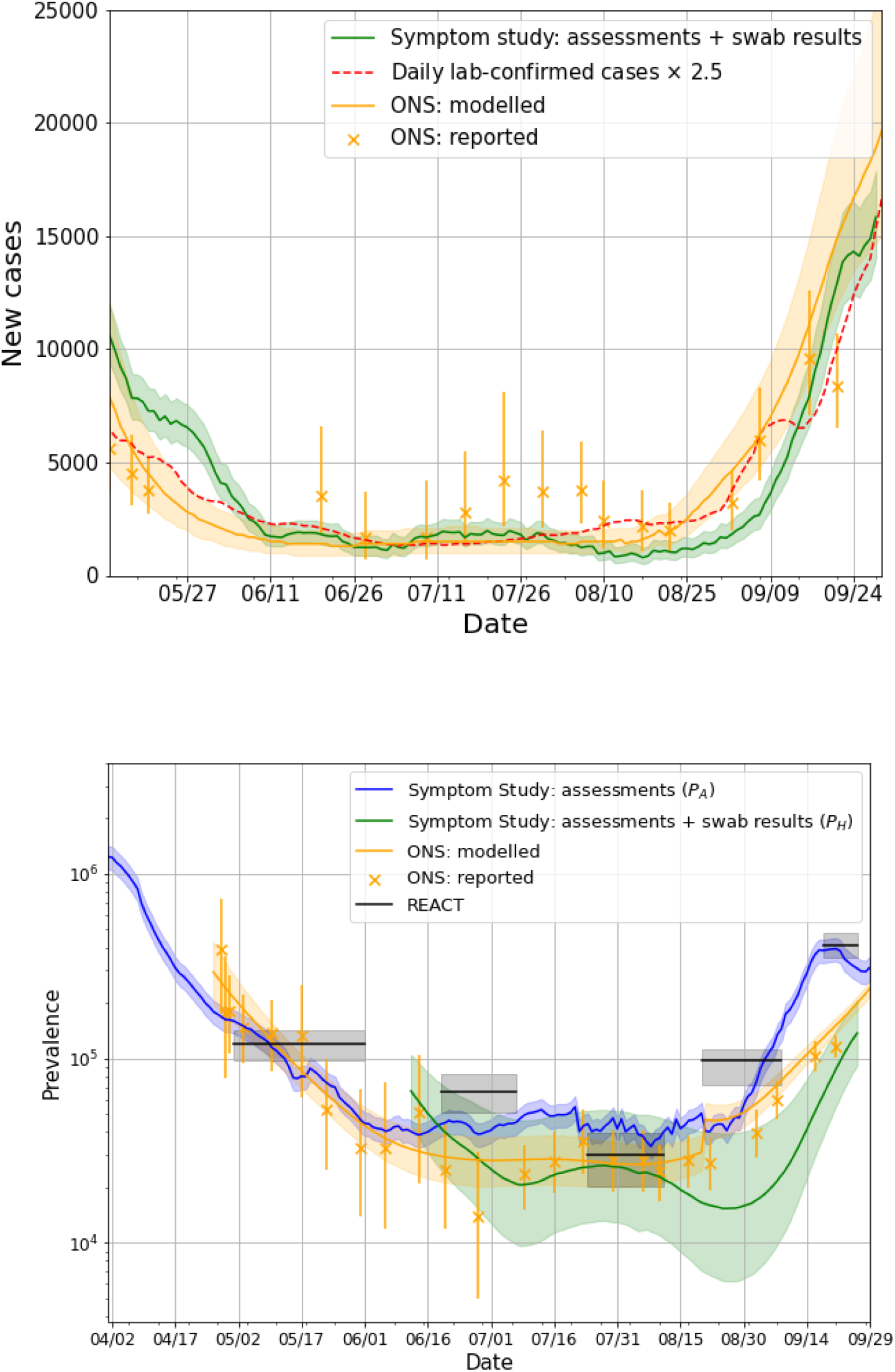
**A)** Daily incidence since 12 May in the UK compared against daily lab-confirmed cases and the ONS study **B)** Daily prevalence in the UK, compared with the ONS and REACT-1 studies. ONS data is taken from the report released on 9 October 2020. ONS report dates are taken as the midpoint for the date range covered by the estimate.

Our results predict a steep decline in incidence until the middle of July, with a trend in agreement with the Government and ONS. All three estimates show an increase in the number of daily cases from mid-August throughout September; on 28 September 2020 we estimated 15,841 (95% CI 14,023-17,885) daily cases. Estimates of incidence per NHS region are shown in Figure S2, and maps of our most granular incidence I_H_ are shown in Figure S3.

Figure 1B compares our England-wide prevalence estimates P_A_ and P_H_ to prevalence reported by the ONS and REACT-1 studies. The app-based assessments P_A_ indicate a continuous drop in the number of cases from 1 April, following the lockdown measures instigated on 23 March, plateauing in mid-June and beginning to rise again sharply from early September. The trends observed for P_A_ agree with data from the ONS survey, which begins on 26 April, and the REACT-1 study, which begins on 1 May. There is some divergence in late September, when both P_A_ and REACT-1 show a sharper rise in prevalence than the ONS study. Note that P_A_ only captures symptomatic cases, whilst the ONS and REACT-1 also capture asymptomatic cases which are thought to account for 40-45% of the total cases^18^ - taking this into account, P_A_ is slightly higher than ONS and REACT-1. On 28 September 2020 we estimated a prevalence of 0.53% (95% CI 0.45-0.60).

P_H,_ agrees with the trends in the other estimates, and predicts a rise in cases in late September at a similar rate to P_A_ and REACT. The absolute values are consistently lower than other estimates. The recovery model used to calculate P_H_ is shown in Figure 2. Whilst most users recover in 7-10 days, the curve shows there is a significant minority who take longer than three weeks to recover from COVID-19.

**Figure 2.**
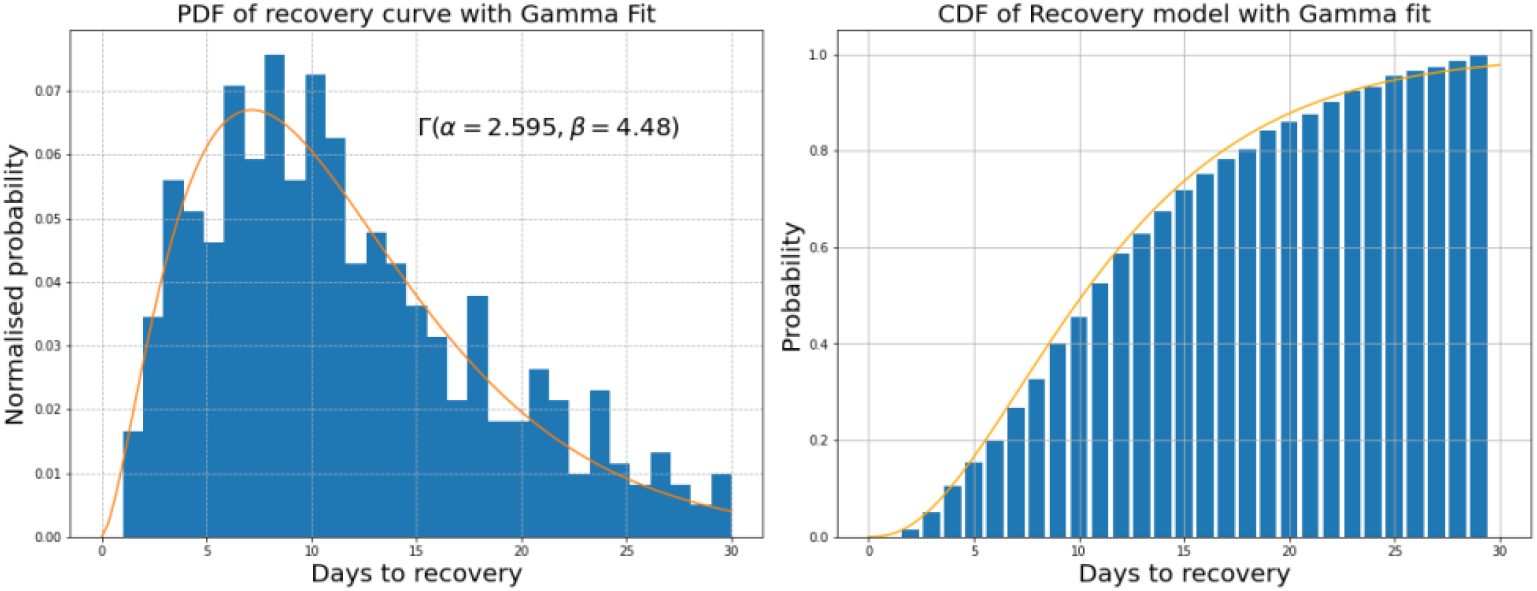
Left: Empirical PDF of days to recovery along with a Gamma fit. Right: Empirical CDF of days to recovery along with the same Gamma fit.

The size of our dataset allows us to estimate prevalence for more granular geographic regions than the ONS (see Supplementary Figure S4). We considered that our estimates of prevalence might be biased by a user’s health-seeking behaviour. We sought to assess the influence of this factor by removing from the analysis all users who reported being sick upon sign-up, Figure S4.

Figure 3 shows estimates of R(t) for each of the NHS regions in England between 24 June and 28 September, compared with the consensus estimates provided by the UK Government’s Science Pandemic Influenza Modelling group (SPI-M)^19^. The estimates both agree that R(t) has been above 1 from early September, we estimate R(t) in England was 1.17 (95% credible interval 1.15-1.19) on 28 September 2020. The Government estimates are much smoother, likely because they are derived from a consensus of the R(t) estimates from the models produced by many groups.

**Figure 3.**
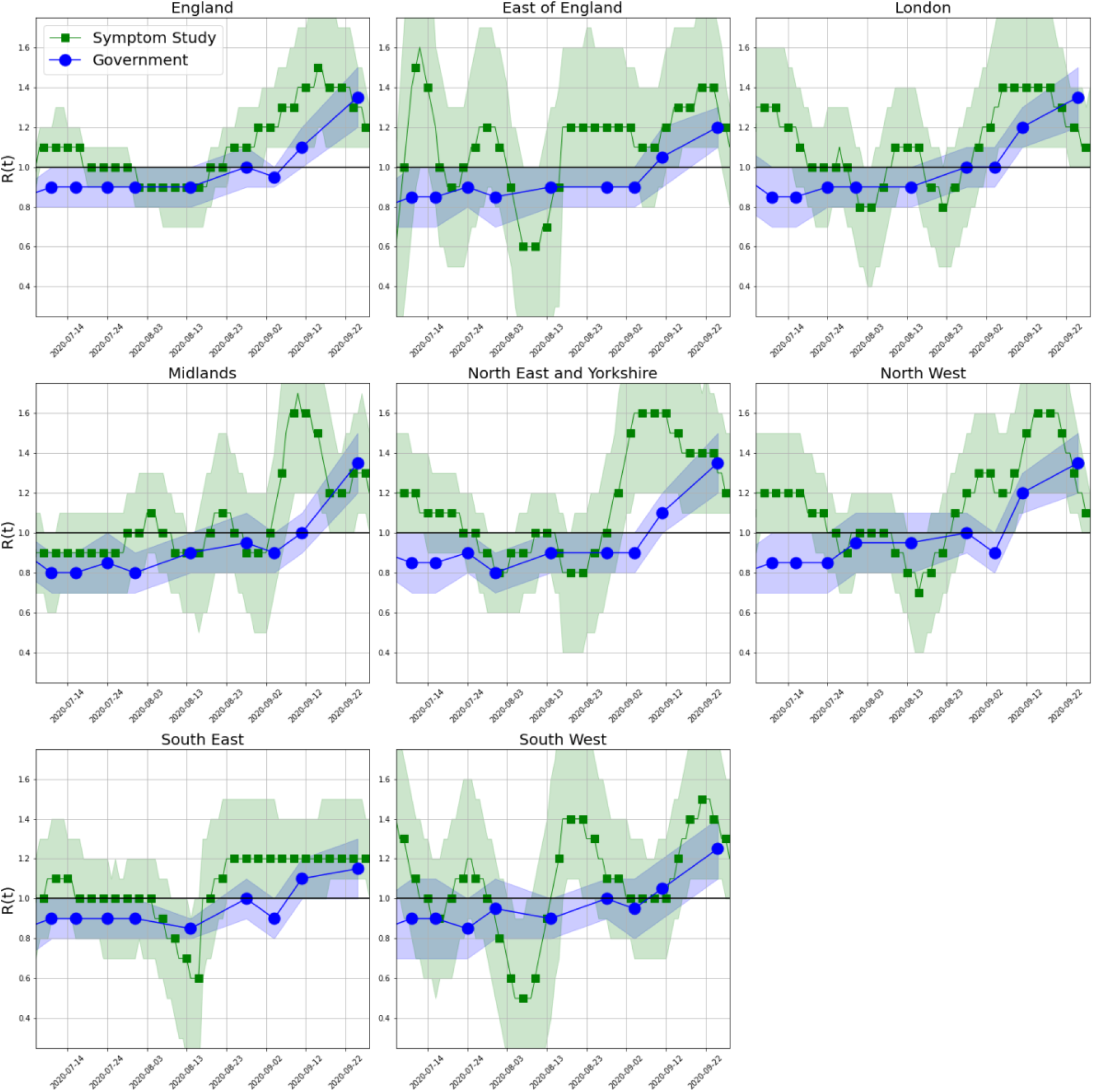
Estimated R(t), for NHS regions in England between 24 June and 28 September, with 95% credible intervals shown. UK government estimates published every 7-12 days from 12 June.

Figure 4 shows the results for hotspot detection in England. Ranking based on incidence consistently outperforms the prevalence-based ranking. Performance varies over time; the best incidence-based ranking produces recall scores of up to 0.80, with a minimum of 0.35, indicating we can predict between 7-16 of the regions in the Government’s top twenty. The ranking performed best in late September when cases began to rise, due to there being greater differences between regional case numbers. Figure 5 compares agreement of weekly cases in each UTLA between Symptom Study and Government numbers against the number of Government Pillar 2 tests carried out. The correlation indicates that the two estimates agree best when the Government carries out more tests. This means that disagreements between the two rankings may be partially explained by poorer ranking in regions with limited Government testing, indicating that our results could provide valuable forecasting in regions with poor testing provision.

**Figure 4.**
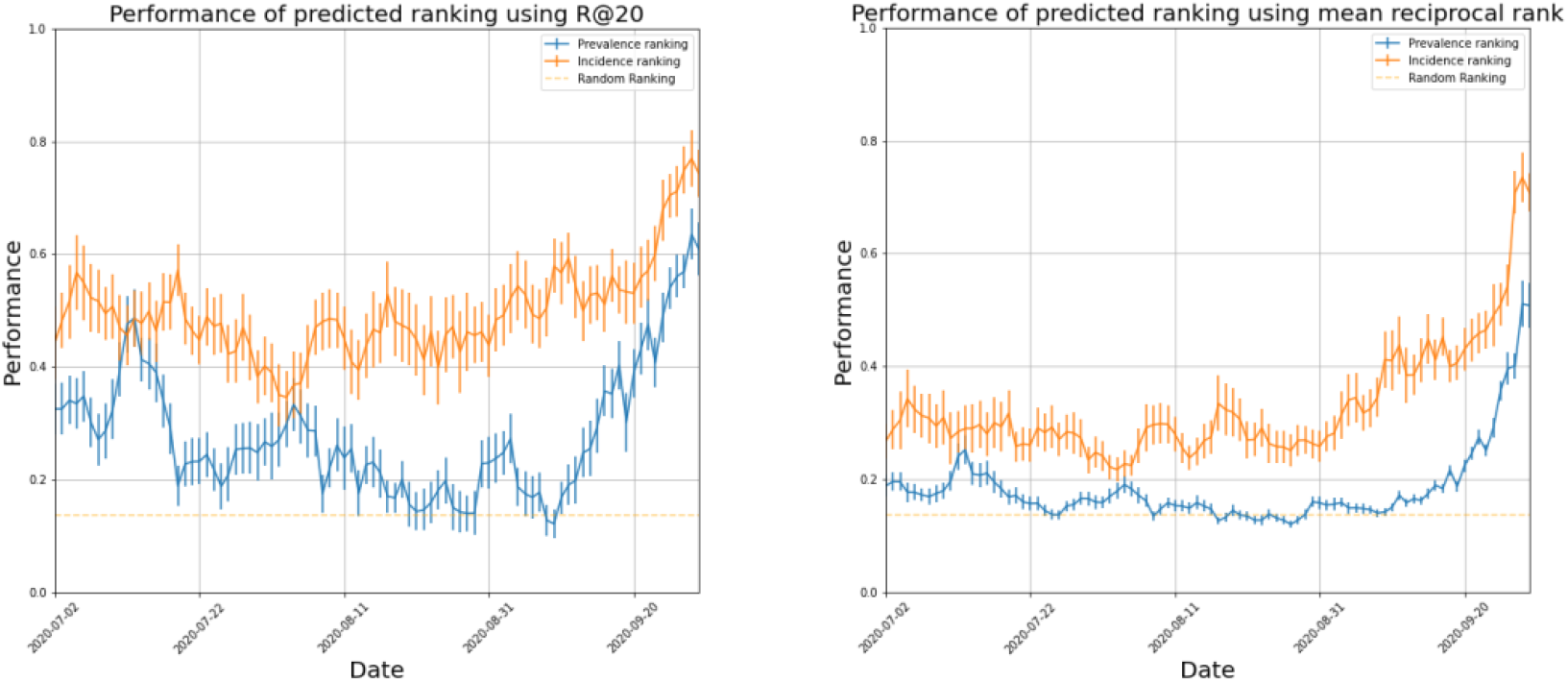
Performance of our two ranking methods: ranking by prevalence and incidence on two metrics, Recall @ 20 and the Normalised Mean Reciprocal Rank.

**Figure 5.**
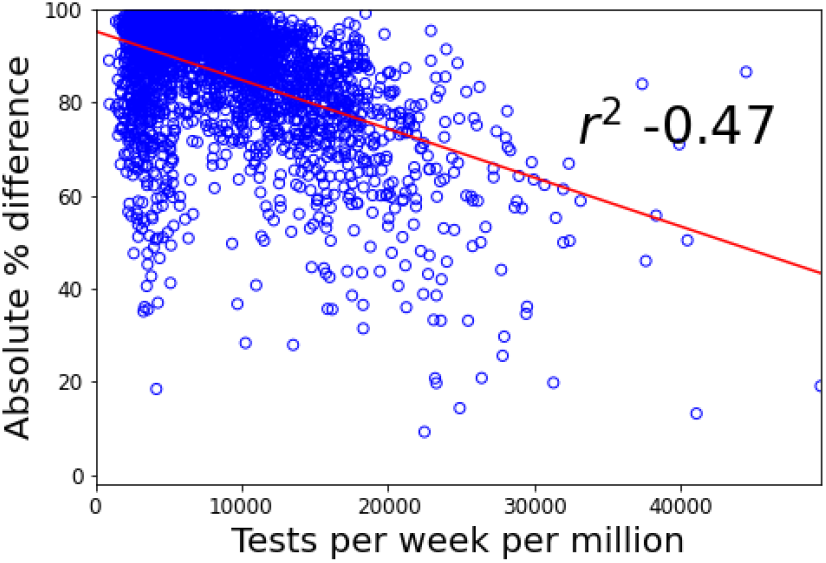
Agreement between Symptom Study and Government case numbers per week and UTLA, against the number of Government Pillar 2 tests carried out.

## Discussion

In this work, we have demonstrated the use of population-wide data reported through the COVID Symptom Study app. Using over 120 million daily reports and 170,000 invited swab tests from 2.8 million users, we were able to estimate prevalence, incidence, and R(t). Other digital surveys have been used to provide valuable real time information about the pandemic. These include How We Feel^20^, Corona Israel^5^, the Facebook Survey, and CovidNearYou. However, to our knowledge we are the first to provide national-level disease surveillance, and find good agreement with traditional, representative community surveys.

Furthermore, we use our data to produce geographically granular estimates and a list of potential hotspots. The list consistently flags a number of regions highlighted by the Government’s testing data. Whilst we compare to data from Government testing in our results, it must be noted these cannot be considered ground truth estimates of COVID-19 cases. The Government data is an incomplete sampling of new COVID-19 cases; results from our app indicate that only 40% of those who report classic COVID-19 symptoms go on to receive a test. Furthermore, testing capacity is not uniform across all UTLAs^21^. Our results indicate that our case estimates agree best with Government estimates in areas where their levels of testing per capita are high, suggesting that our estimates could prove a valuable resource in regions with limited testing. There are other reasons our list may differ from the Government’s: the two methods may have different uptake in certain higher-risk groups, such as students in provided accommodation, thus showing different sensitivity to hotspots based on the demographic make-up of a region. The two methods may therefore be complementary and we suggest our hotspot detection may be most beneficial as an additional indication of regions where increased testing might be best focused. The modest reliance on PCR tests suggests our approach could prove valuable in countries where testing infrastructure is more limited, though further work is required to assess our approach in other locations.

Other efforts to track the national progression of COVID-19 rely on self-swabbing from community cohorts. Two such efforts exist in England: the ONS study^17^ and the REACT-1 study^8,9^. These studies have the advantage of being more representative of the population, and their design enables the detection of asymptomatic cases. However, they are smaller than the Covid Symptom Study: the ONS and REACT-1 currently report between 120,000-175,000 participants in England, whilst the app reports over 2,800,000 users in England. The ability to use self-reported symptom data from this large cohort enables us to make predictions of more geographically granular regions than either the ONS or REACT-1 studies, enabling us to predict COVID-19 hotspots at the UTLA level. Our estimates should thus be viewed as independent and complementary to those provided by the ONS and REACT-1 studies.

Several limitations to our work must be acknowledged. The app users are not a representative sample of the wider population for which we aim to make an inference. There is a clear shift in age and gender compared to the general population, our users tend to live in less deprived area^12^, and we have few users reporting from key sites such as care homes and hospitals. We account for some population differences when producing prevalence estimates using specific census adjusted population strata, but the number of invited tests do not allow us to do this when calculating incidence. Differences in reported symptoms across age groups^23^ would likely lead to different prediction models of COVID-19 positivity, and the model’s performance will vary with the prevalence of other infections with symptoms that overlap with COVID-19, such as flu. Furthermore, the app population is less racially and ethnically diverse than the general population.^24^. Reliance on user self-reporting can also introduce bias into our results - for instance, users who are very sick may be less likely to report than those with mild symptoms. Other sources of error include collider bias^25^ arising from a user’s probability of using the app being dependent on their likelihood of having COVID-19, potentially biasing our estimates of incidence and prevalence. We showed a sensitivity analysis that attempts to understand the effect of health-seeking behaviour, but acknowledge there are many other biases that may affect our results - for example our users may be more risk-averse than the general population- and that our results must be interpreted with this in mind.

## Conclusion

We have presented a means of combining app-based symptom reports and targeted testing from over 2.8 million users to estimate incidence, prevalence and R(t) in England. By integrating symptom-reports with PCR test results we were able to highlight regions which may have concerning increases in COVID-19 cases. This approach could be an effective, complementary way for Governments to monitor the spread of COVID-19 and identify potential areas of concern.

## Ethics

Ethics has been approved by KCL Ethics Committee REMAS ID 18210, review reference LRS-19/20-18210 and all participants provided consent.

## Data sharing

Data collected in the COVID-19 Symptom Study smartphone application are being shared with other health researchers through the UK National Health Service-funded Health Data Research UK (HDRUK) and Secure Anonymised Information Linkage consortium, housed in the UK Secure Research Platform (Swansea, UK). Anonymised data are available to be shared with HDRUK researchers according to their protocols in the public interest (https://web.www.healthdatagateway.org/dataset/fddcb382-3051-4394-8436-b92295f14259). US investigators are encouraged to coordinate data requests through the Coronavirus Pandemic Epidemiology Consortium (https://www.monganinstitute.org/cope-consortium).

## Author Contributions

TV, MSG, JW, SO, CJS, TDS, TF, MG, PWF contributed to study concept and design. SG, JCP, CHS, DAD, LHN, ATC, RD, JW, CJS, TDS, SO contributed to acquisition of data. TV, MSG, LSC, SG, JCP, CHS, BM contributed to data analysis and have verified the underlying data. TV, MSG, LSC contributed to initial drafting of the manuscript. All authors contributed to interpretation of data and critical revision of the manuscript. ATC, CJS, TDS, SO contributed to study supervision.

## Declaration of interests

JW, RD, JCP, SG are employees of Zoe Global Ltd. TDS is a consultant to Zoe Global Ltd. DAD and ATC previously served as investigators on a clinical trial of diet and lifestyle using a separate smartphone application that was supported by Zoe Global. MFG reports non-financial support and other from Novo Nordisk, non-financial support and other from Pfizer, non-financial support and other from Follicum, non-financial support and other from Abcentra, non-financial support from Probi, non-financial support from Johnson & Johnson, grants from EU H2020-JTI-lMl2-2015-05 (Grant agreement number 115974 - BEAt-DKD), non-financial support and other from Boehringer Ingelheim Pharma GmbH, non-financial support and other from JDRF International, non-financial support and other from Eli Lilly, non-financial support and other from AbbVie, non-financial support and other from Sanofi-Aventis, non-financial support and other from Astellas, personal fees from Lilly, non-financial support and other from Novo Nordisk A/S, non-financial support and other from Bayer AG, outside the submitted work. ATC reports grants from Massachusetts Consortium on Pathogen Readiness, during the conduct of the study; personal fees from Pfizer Inc., grants and personal fees from Bayer Pharma AG, personal fees from Boehringer Ingelheim, outside the submitted work; TF reports grants from ERC, grants from Swedish Research Council, grants from FORTE Research Council, grants from Swedish Heart & Lund foundation, outside the submitted work; All other authors declare no competing interests.

## Acknowledgements

Zoe provided in kind support for all aspects of building, running and supporting the app and service to all users worldwide. Support for this study was provided by the NIHR-funded Biomedical Research Centre based at GSTT NHS Foundation Trust. Investigators also received support from the Wellcome Trust, the MRC/BHF, Alzheimer’s Society, EU, NIHR, CDRF, and the NIHR-funded BioResource, Clinical Research Facility and BRC based at GSTT NHS Foundation Trust in partnership with KCL, the UK Research and Innovation London Medical Imaging & Artificial Intelligence Centre for Value Based Healthcare, the Wellcome Flagship Programme (WT213038/Z/18/Z), the Chronic Disease Research Foundation, and DHSC. LHN is supported by the American Gastroenterological Association Research Scholars Award and the National Institute of Diabetes and Digestive and Kidney Diseases (K23 DK125838). DAD is supported by the National Institute of Diabetes and Digestive and Kidney Diseases K01DK120742. LHN and DAD are supported by the American Gastroenterological Association AGA-Takeda COVID-19 Rapid Response Research Award (AGA2021-5102). CMA is supported by the National Institute of Diabetes and Digestive and Kidney Diseases (K23 DK120899) ATC was supported in this work through a Stuart and Suzanne Steele MGH Research Scholar Award.The Massachusetts Consortium on Pathogen Readiness (MassCPR) and Mark and Lisa Schwartz supported MGH investigators (LHN, DAD, ADJ, ATC). Investigators from the COVID Symptom Study Sweden were funded in part by grants from the Swedish Research Council, Swedish Heart-Lung Foundation and the Swedish Foundation for Strategic Research (LUDC-IRC 15-0067). TF holds an ERC Starting Grant. Special thank you to Catherine Burrows.

## Supplementary Appendix

### Supplementary Methods

#### Incidence

We use Bayes rule to estimate the number of symptomatic COVID-19 cases in the general population,

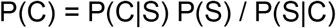

P(C) is the probability that a randomly selected person is infected with COVID-19 on a given day. P(S) is the probability of being newly sick according to the data entered in the app, defined as somebody who logs as healthy for at least nine days before reporting any of the symptoms asked about in the app (appendix pp 3-6). These newly sick users are invited to take a swab test. P(C|S) is the probability of a user testing COVID-19 positive given they are newly sick on the app. This is estimated as the percentage testing positive amongst the newly sick users that accept the test invite, with the assumption that the number of positive cases in this population is representative of the full population of newly sick users. This test positivity rate is considered as a binomial proportion, for which the 95% confidence intervals are derived using the Wilson score.The limits of this confidence interval are substituted in the same conditional probability equation to get the confidence interval for p(C) and in turn for incidence. P(S|C) is the probability of developing symptoms given that one has COVID-19. We set P(S|C) = 1 as we focus on the prediction of symptomatic cases. Note the calculation of p(C) reflects a simplifying assumption that a person becomes COVID-19 positive on the day they first report symptoms, in reality the start of infection will be before this.

#### Symptom-based classifier

The symptom-based classifier used in this work was developed and described in Menni et. al., Nature Medicine (2020). In brief, data from the COVID Symptom Study was used to generate a symptom-based classifier among a subset of 6,452 COVID-19-positive cases and 9,186 COVID-19-negative controls to generate a linear model for likelihood of COVID-19 infection. The model was divided into a train and testing set on a 80:20 ratio. Predictive features were chosen on the basis of a stepwise linear regression, to produce the following model:

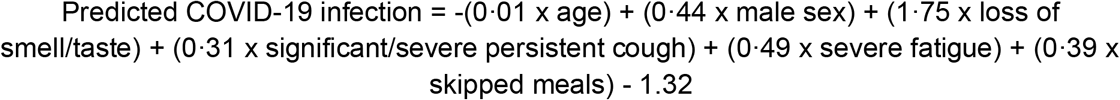

where all symptoms are coded as 1 if the person self-reports the symptom and 0 if not. The sex feature is also binary, with 1 indicative of male participants and 0 representing females. The obtained value is then transformed into predicted probability using exp(x)/(1 + exp(*x*)) transformation followed by assigning cases of predicted COVID-19 for probabilities >0.5 and controls for probabilities <0.5.

In the UK test set, the prediction model had a sensitivity of 0.65 (95% CI 0.62–0.67), a specificity of 0.78 (95% CI0.76–0.80), an area under the curve (AUC) of the receiver operating characteristic curve (ROC) (that is, ROC-AUC) of 0.76 (95% CI 0.74–0.78), a positive predictive value of 0.69 (95% CI 0.66–0.71) and a negative predictive value of 0.75 (95% CI 0.73–0.77). A cross-validation ROC-AUC was 0.75 (95% CI 0.74–0.76) in the 15,638 UK users who were tested for SARS-CoV-2.

#### Model of recovery

The model of recovery, M_t_, describes the proportion of users who become infected on day 0 who will recover on day t. We estimate the model from our data. We looked at users who reported a positive RT-PCR test and were also predicted positive from our symptom-based model. We defined onset as the first appearance of any symptom that occurred less than seven days before a positive test or being predicted positive. We defined recovery as either seven days of uninterrupted healthy reporting in the app or the date of a negative RT-PCR test, selecting the smallest value if both occurred. For our prevalence estimate, we truncate the model of recovery at 30 days, meaning we consider anyone who has suffered from COVID-19 for greater than 30 days as a long-term COVID-19 patient who is no longer infectious. This agrees with (Wajnberg et. al., 2020) where the authors suggest that although patients can test positive on RT-PCR beyond 28 days, they are unlikely to be infectious. This gives a probability distribution for the number of days it takes to recover from COVID-19.

Using the incidence estimates per NHS region, I_S_, we can produce prevalence estimates per NHS region using the following dot-product:

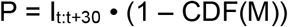

Where I_t:t+30_ is a vector of incidence estimates in the 30 days up to time t and CDF(M) is the cumulative distribution function of the model of recovery, M.

### Supplementary Tables

**Table S1.** Questions asked in the COVID Symptom Study App. The application is available to users in both private and non-private accommodation (e.g. care homes, student accommodation). Users are asked to enter their location on sign-up, and there is an option to update the location if a user moves.

| Baseline questions (asked on sign-up) |  |
| --- | --- |
| About your work |  |
| Are you a health care worker (including hospital, elderly care or in the community)? | - Yes, currently treat patients<br>- Yes, do not currently treat patients<br>- Yes, I currently interact with patients<br>- Yes, but I do not currently interact with patients<br>- No |
| Do you care for multiple people in the community, with direct contact with your patients? | - Yes/No |
| **About you** |  |
| What year were you born? |  |
| What sex were you assigned at birth? | - Female<br>- Male<br>- Prefer not to say<br>- Intersex |
| What gender do you most identify with? | - Male<br>- Female<br>- Transgender<br>- Do not identify as female, male or transgender<br>- Prefer not to say |
| Which of the following best describes your ethnicity? | - Asian/Asian British - Indian, Pakistani, Bangladeshi, other<br>- Black/Black British - Caribbean, African, other<br>- Mixed race - White and Black/Black British<br>- Mixed race - other<br>- White - British, Irish, other<br>- Chinese/Chinese British<br>- Middle Eastern/Middle Eastern British - Arab, Turkish, other<br>- Other ethnic group<br>- Prefer not to say |
| What is your height? |  |
| What is your weight? |  |
| What is your postcode? |  |
| In general, do you have any health problems that require you to stay at home? | - Yes/No |
| **About your health** |  |
| Do you have heart disease? | - Yes/No |
| Do you have heart disease? | - Yes/No |
| Do you have lung disease or asthma? | - Yes/No |
| Do you smoke? | -Yes<br>- Not currently<br>- Never |
| Do you have kidney disease? | Yes/No |
| Are you living with cancer? | Yes/No |
| **Follow up questions (asked daily)** |  |
| **Tests** |  |
| Have you had a test for COVID-19? | Yes/No |
| Did you test positive for COVID-19? | - Positive<br>- Negative<br>- Test failed<br>- Waiting for result |
| How was this test performed? | - Nose swab<br>- Throat swab<br>- Spit tube<br>- Blood sample |
| When was your test? |  |
| **Symptoms** |  |
| How do you feel right now? | - I feel as healthy as normal<br>- I'm not feeling quite right |
| If not feeling quite right: |  |
| Do you have a fever? | Yes/No |
| Do you feel chills or shivers (feel too cold)? | - Yes/No |
| If you are able to measure it, what is your temperature? |  |
| Do you have a persistent cough (coughing a lot for more than an hour, or 3 or more coughing episodes in 24 hours)? | - Yes/No |
| Are you experiencing unusual fatigue? | - No<br>- Mild fatigue<br>- Severe fatigue - I struggle to get out of bed |
| Are you experiencing unusual shortness of breath? | - No<br>- Yes. Mild symptoms - slight shortness of breath during ordinary activity<br>- Yes. Significant symptoms - breathing is comfortable only at rest<br>- Yes. Severe symptoms - breathing is difficult even at rest" |
| Do you have a loss of smell/taste? | Yes/No |
| Do you have an unusually hoarse voice? | Yes/No |
| Are you feeling an unusual chest pain or tightness in your chest? | Yes/No |
| Do you have an unusual abdominal pain? | Yes/No |
| Are you experiencing diarrhoea? | Yes/No |
| Do you have a headache? | - Yes/No |
| Do you have any of the following symptoms: confusion, disorientation or drowsiness? | Yes/No |
| Do your eyes have any unusual eye-soreness or discomfort (e.g. light sensitivity, excessive tears, or pink/red eye)? | Yes/No |
| Have you been skipping meals? | Yes/No |
| Are you experiencing dizziness or light-headedness? | Yes/No |
| Do you have a sore throat? | Yes/No |
| Do you have unusual strong muscle pains? | Yes/No |
| Have you had raised, red, itchy welts on the skin or sudden swelling of the face or lips? | Yes/No |
| Have you had any red/purple sores or blisters on your feet, including your toes? | Yes/No |
| Are there other important symptoms you want to share with us? | Free text entry |

### Supplementary Figures

**Figure S1.**
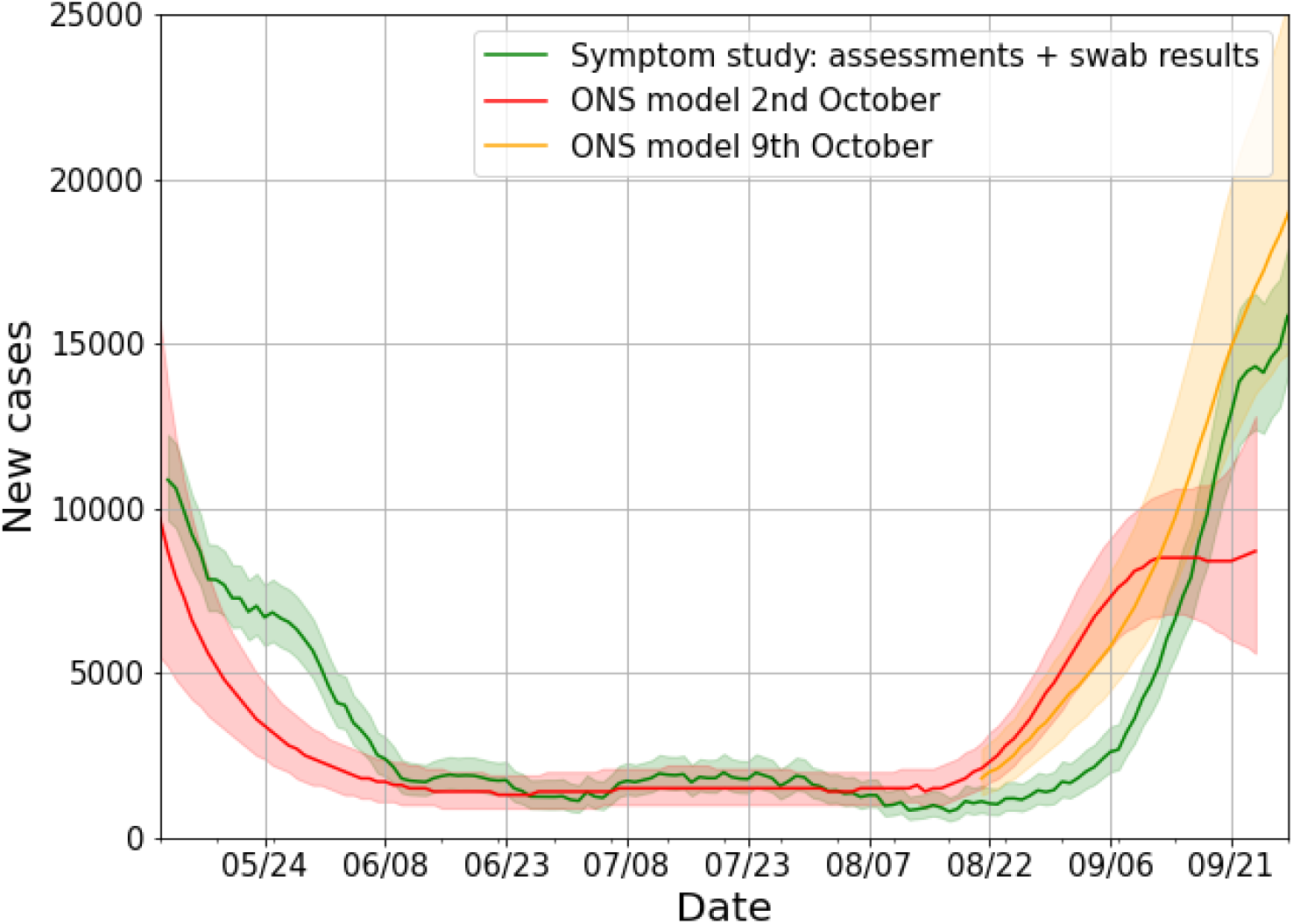
Comparison of incidence data with two ONS models. Comparison of our incidence data with two ONS models, released on 2 October and 9 October. The model released on 2 October showed incidence levelling off in September, in disagreement with our released incidence estimates. By contrast the 9 October model showed a rapid increase in daily cases throughout September.

**Figure S2.**
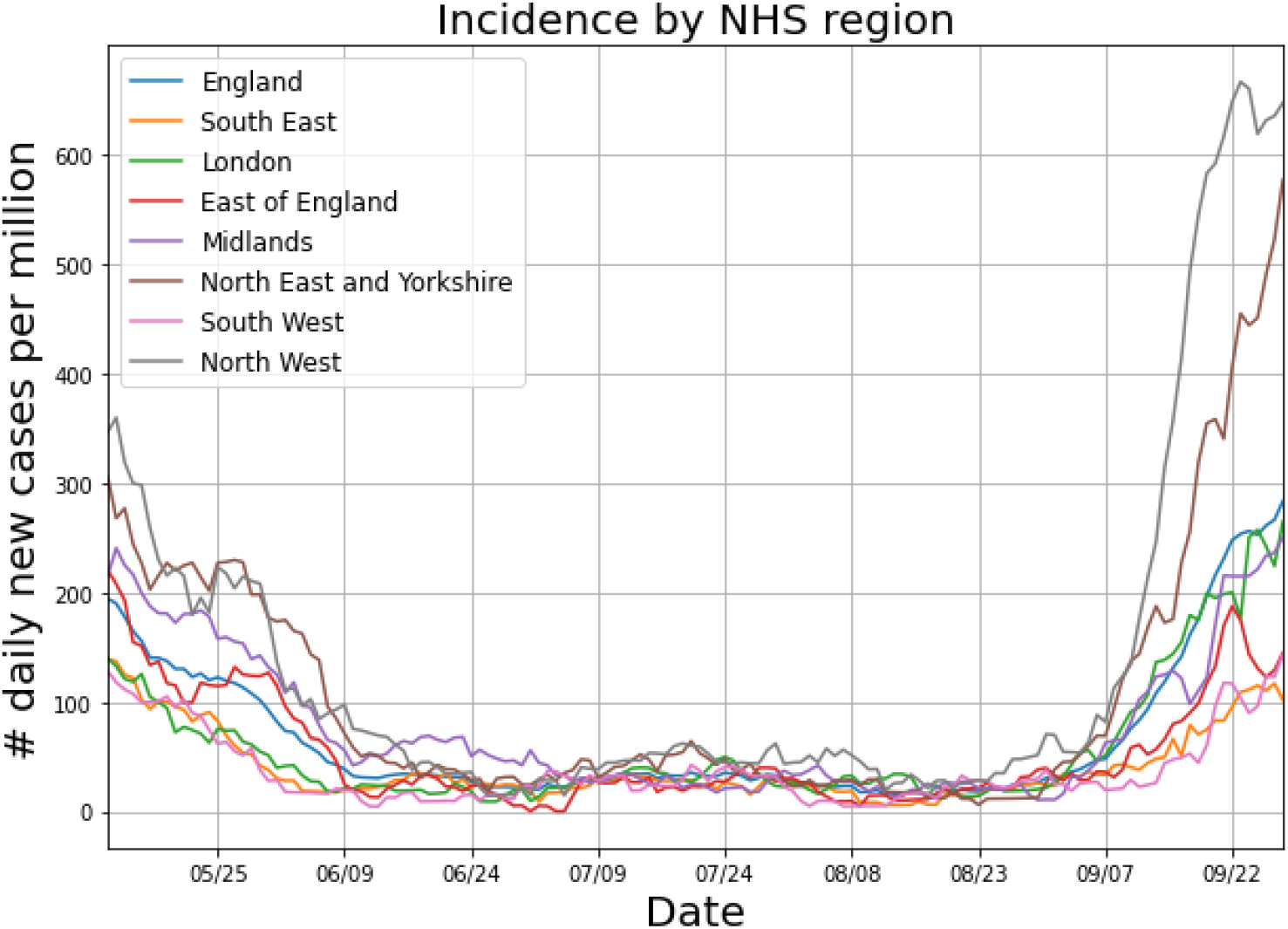
Estimated incidence for each NHS region in England.

**Figure S3.**
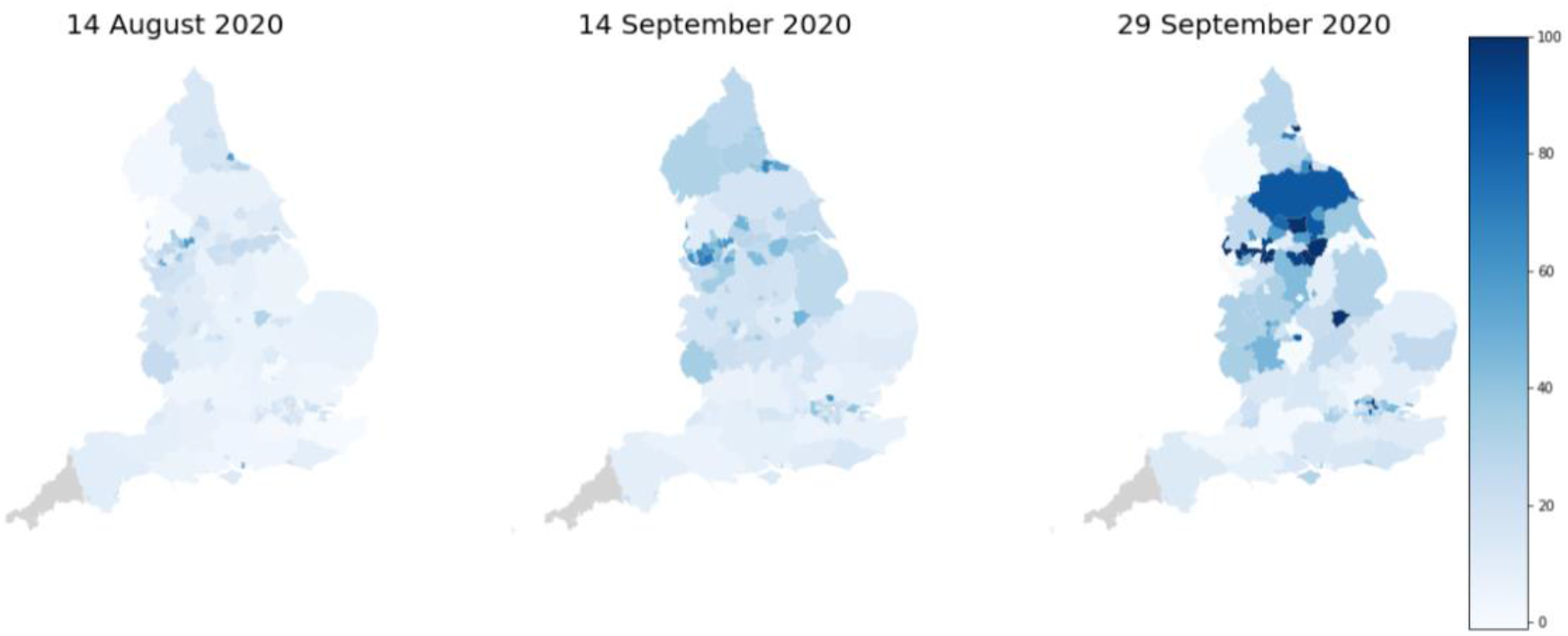
Map of daily cases per 100,000 people per UTLA. Maps show daily cases per 100,000 people in each UTLA at three time-points, selected to show the lowest number of cases in mid-August, and the rapid increase through September.

**Figure S4.**
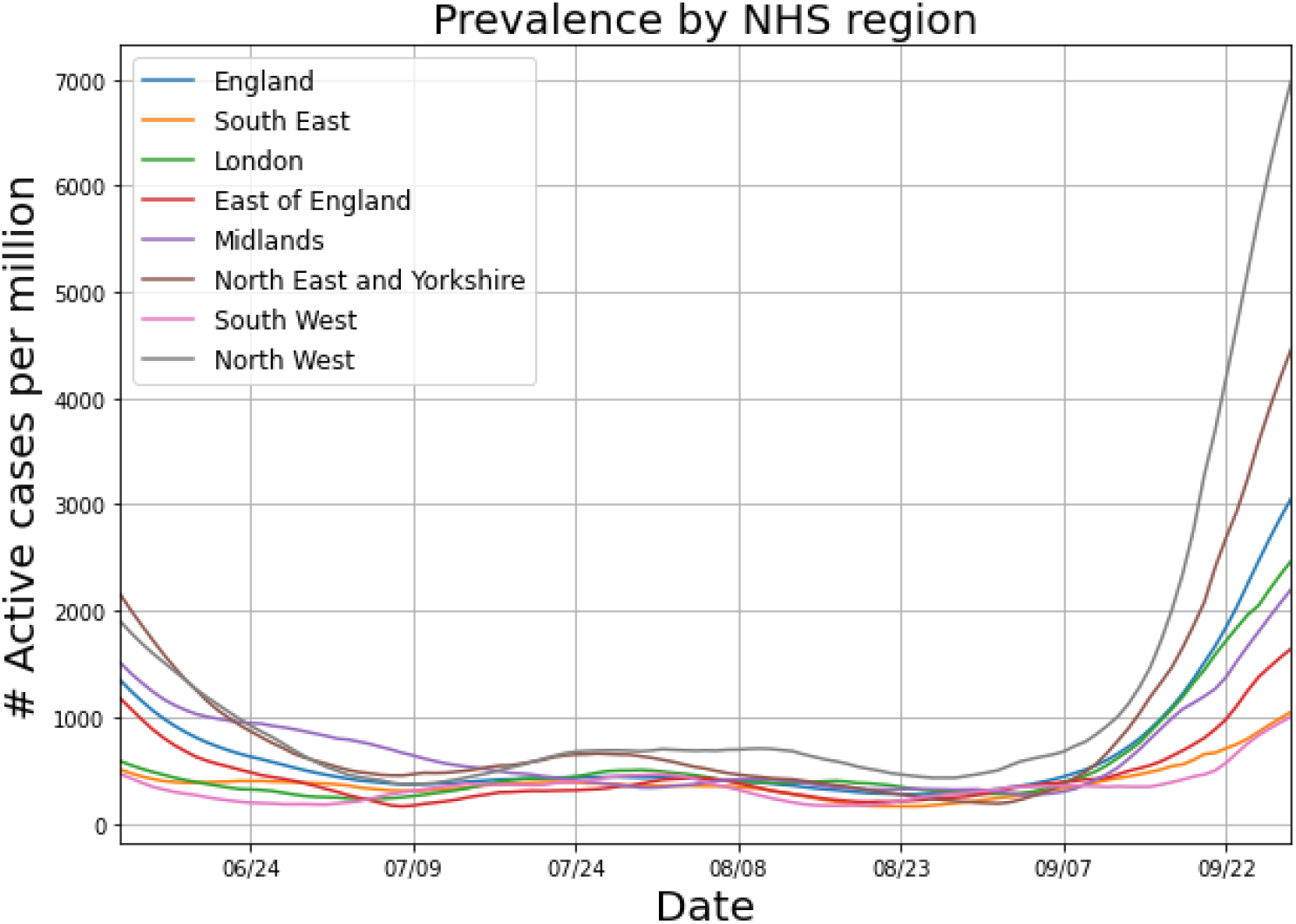
Estimated prevalence for each NHS region in England.

**Figure S5.**
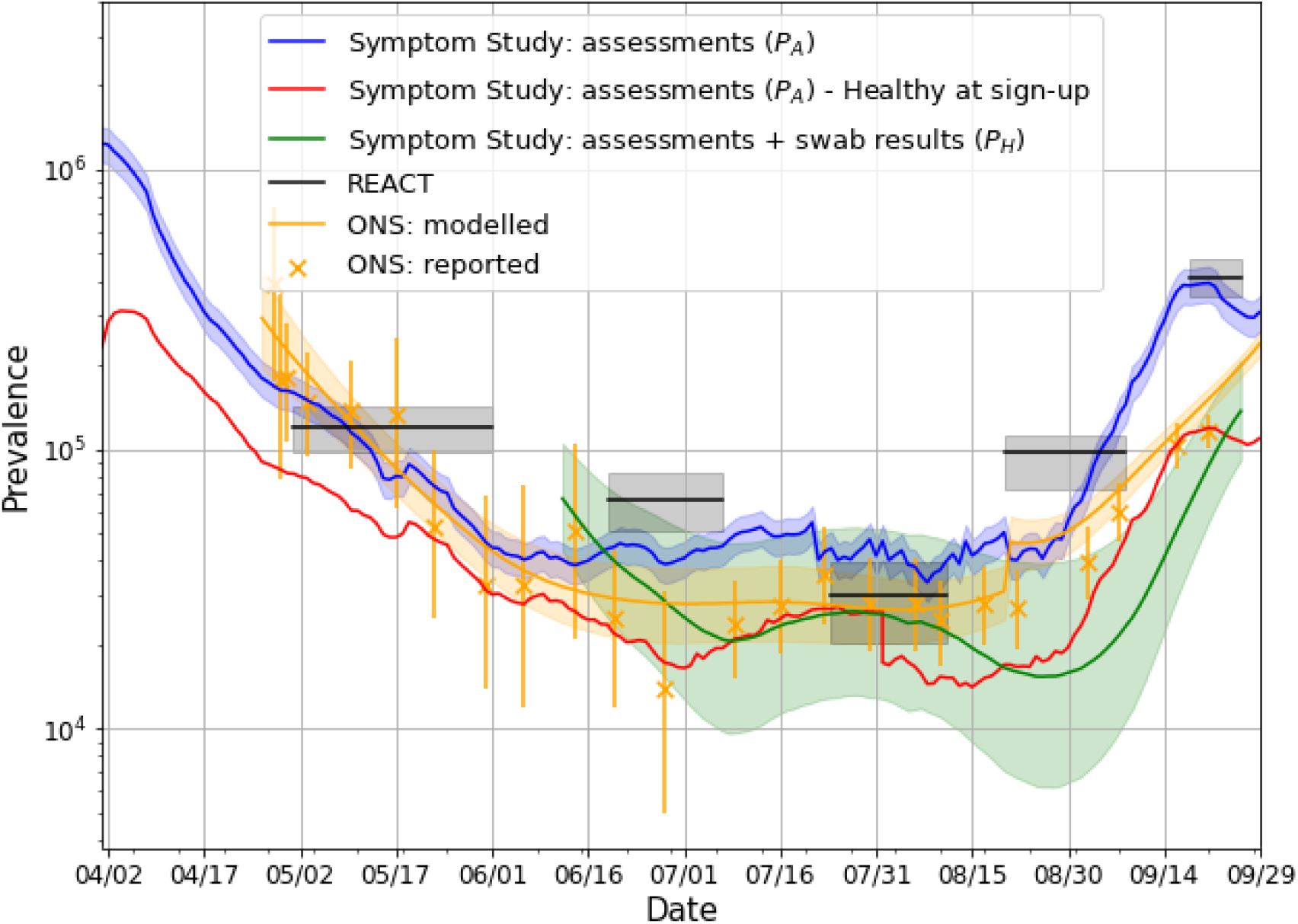
Estimates of prevalence, including an estimate obtained when all users who are sick upon sign-up are dropped from the analysis.

**Figure S6.**
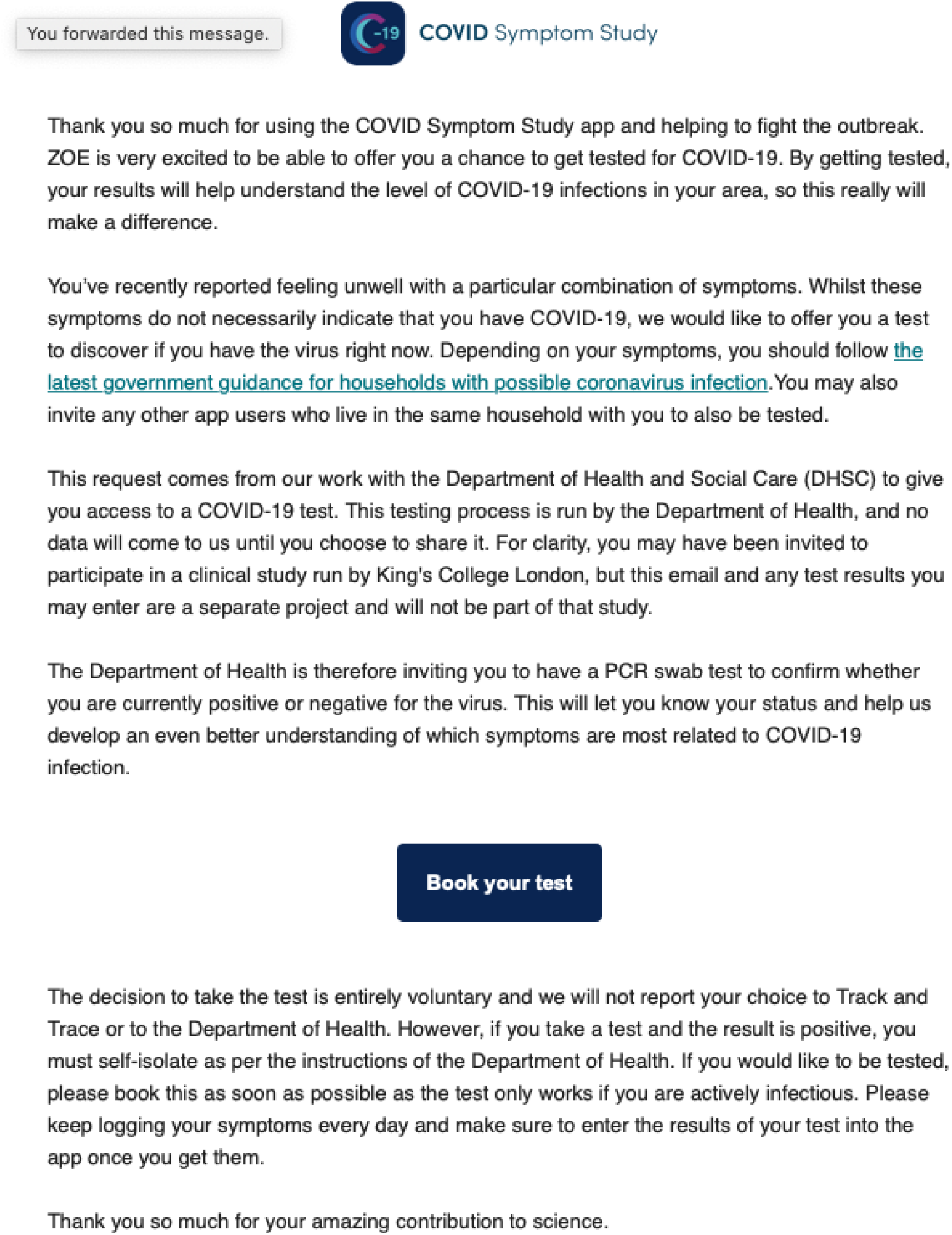
The testing invite sent to app users.

## Notes

### Clinical Trial

NCT04331509

### Summary of Updates

Paper updated in response to peer review.

## References

1 Flaxman S, Mishra S, Gandy A, et al. Estimating the effects of non-pharmaceutical interventions on COVID-19 in Europe. Nature 2020; 584 : 257–61.

2 Mahase E. Covid-19: How does local lockdown work, and is it effective? 2020. https://www.bmj.com/content/370/bmj.m2679.abstract.

3 Lavezzo E, Franchin E, Ciavarella C, et al. Suppression of a SARS-CoV-2 outbreak in the Italian municipality of Vo’. Nature 2020; 584 : 425–9.

4 Peto J, Alwan NA, Godfrey KM, et al. Universal weekly testing as the UK COVID-19 lockdown exit strategy. Lance t 2020; 395 : 1420–1.

5 Rossman H, Keshet A, Shilo S, et al. A framework for identifying regional outbreak and spread of COVID-19 from one-minute population-wide surveys. Nat Med 2020; 26 : 634– 8.

6 Drew DA, Nguyen LH, Steves CJ, et al. Rapid implementation of mobile technology for real-time epidemiology of COVID-19. Science 2020; published online May 5. DOI: 10.1126/science.abc0473.

7 Pouwels KB, House T, Pritchard E, et al. Community prevalence of SARS-CoV-2 in England during April to September 2020: Results from the ONS Coronavirus Infection Survey. MedRxiv 2020. https://www.medrxiv.org/content/10.1101/2020.10.26.20219428v1.abstract.

8 Riley S, Atchison C, Ashby D, et al. REal-time Assessment of Community Transmission (REACT) of SARS-CoV-2 virus: Study protocol. Wellcome Open Research 2020; 5 : 200.

9 Riley S, Ainslie KEC, Eales O, Walters CE, Wang H. High prevalence of SARS-CoV-2 swab positivity in England during September 2020: interim report of round 5 of REACT- 1 study. medRxiv 2020. https://www.medrxiv.org/content/10.1101/2020.09.30.20204727v1.abstract.

10 Government Testing Methodology. https://www.gov.uk/government/publications/coronavirus-covid-19-testing-data-methodology/covid-19-testing-data-methodology-note (accessed Sept 11, 2020).

11 Menni C, Valdes AM, Freidin MB, et al. Real-time tracking of self-reported symptoms to predict potential COVID-19. Nat Med 2020; published online May 11. DOI: 10.1038/s41591-020-0916-2.

12 Bowyer R, Varsavsky T, Sudre CH, et al. Geo-social gradients in predicted COVID-19 prevalence and severity in Great Britain: results from 2,266,235 users of the COVID-19 Symptoms Tracker app. Epidemiology. 2020; published online April 27. DOI: 10.1101/2020.04.23.20076521.

13 McLennan D, Noble S, Noble M, Plunkett E, Wright G, Gutacker N. The English Indices of Deprivation 2019: technical report. 2019. https://dera.ioe.ac.uk/34259/1/IoD2019_Technical_Report.pdf.

14 Bettencourt LMA, Ribeiro RM. Real time bayesian estimation of the epidemic potential of emerging infectious diseases. PLoS One 2008; 3 : e2185.

15 Nishiura H, Linton NM, Akhmetzhanov AR. Serial interval of novel coronavirus (COVID- 19) infections. Int J Infect Dis 2020; 93 : 284–6.

16 Murray B, Kerfoot E, Graham MS, et al. Accessible Data Curation and Analytics for International-Scale Citizen Science Datasets. arXiv [cs.DB]. 2020; published online Nov 2. http://arxiv.org/abs/2011.00867.

17 Pouwels KB, House T, Robotham JV, Birrell P. Community prevalence of sars-cov-2 in england: Results from the ons coronavirus infection survey pilot. medRxiv 2020. https://www.medrxiv.org/content/10.1101/2020.07.06.20147348v1.abstract.

18 Oran DP, Topol EJ. Prevalence of Asymptomatic SARS-CoV-2 Infection : A Narrative Review. Ann Intern Med 2020; 173 : 362–7.

19 UK Government Published R Estimates. https://www.gov.uk/guidance/the-r-number-in-the-uk. (accessed Sept 30, 2020).

20 Allen WE, Altae-Tran H, Briggs J, et al. Population-scale longitudinal mapping of COVID-19 symptoms, behaviour and testing. Nat Hum Behav 2020; 4 : 972–82.

21 Medicine TLR, The Lancet Respiratory Medicine. COVID-19 testing in the UK. The Lancet Respiratory Medicine. 2020; 8 : 1061.

22 Government testing dashboard. https://coronavirus.data.gov.uk/testing (accessed Oct 20, 2020).

23 Zazzara MB, Penfold RS, Roberts AL, et al. Probable delirium is a presenting symptom of COVID-19 in frail, older adults: a cohort study of 322 hospitalised and 535 community-based older adults. Age and Ageing. 2020. DOI: 10.1093/ageing/afaa223.

24 Lo C-H, Nguyen LH, Drew DA, et al. Racial and ethnic determinants of Covid-19 risk. medRxiv 2020. https://www.medrxiv.org/content/10.1101/2020.06.18.20134742v1?rss=1%22.

25 Griffith G, Morris TT, Tudball M, et al. Collider bias undermines our understanding of COVID-19 disease risk and severity. medRxiv 2020. https://www.medrxiv.org/content/10.1101/2020.05.04.20090506v3.abstract.

